# Factors associated with access to virtual care in older adults: A cross-sectional study

**DOI:** 10.1101/2020.10.01.20205302

**Authors:** Laura Liu, Zahra Goodarzi, Aaron Jones, Ron Posno, Sharon Straus, Jennifer Watt

**Author notes:** Correspondence: Dr. Jennifer Watt, St. Michael’s Hospital, 4-002 Shuter Wing, 30 Bond St, Toronto ON M5B 1W8.

## Abstract

**Background:** During the COVID-19 pandemic, virtual care (i.e. telephone or videoconference) has played a critical role. However, concerns were raised regarding equitable access for older adults, in particular, given potential advantages of videoconference-as opposed to telephone-based assessment. Our objective was to describe patient-specific factors associated with different modes of virtual healthcare.

**Methods:** We reviewed medical records of all patients assessed virtually in the geriatric medicine clinic at St. Michael’s Hospital, Toronto, Canada, between March 17 and July 13, 2020. We derived adjusted odds ratios (OR), risk differences (RD), and marginal and predicted probabilities, with 95% confidence intervals, from a multivariable logistic regression model, which tested the association between having a videoconference assessment (vs. a telephone assessment) and patient age, sex, ability to use a computer, education, frailty (measured on the Clinical Frailty Scale), history of cognitive impairment, and immigration history; language of assessment, and caregiver involvement in assessment.

**Results:** Our study included 330 patients (227 telephone and 103 videoconference assessments). Frailty (adjusted OR 0.62, 0.45 to 0.85; adjusted RD -0.08, -0.09 to -0.06) and absence of a caregiver (adjusted OR 0.12, 0.06 to 0.24; adjusted RD -0.35, -0.43 to -0.26) were associated with lower odds of videoconference assessment. For example, an 80-year-old woman with mild frailty who immigrated to Canada, speaks English, attained a post-secondary education, does not have cognitive impairment, and uses a computer had a 60% (39% to 80%) predicted probability of videoconference assessment if a caregiver was present compared to 15% (3% to 26%) without a caregiver. Only 32 of 98 (32.7%) patients who could independently use a computer participated in videoconference assessments.

**Conclusion:** Given the recent expansion of virtual care, we must urgently implement and evaluate strategies that optimise equitable access to videoconference-based virtual care for older adults.

## Introduction

Virtual care (i.e. videoconference or telephone) has rapidly expanded to meet patient needs and physical distancing guidelines during the COVID-19 pandemic.^1-3^ Virtual care provides a way for healthcare providers to interact with patients without needing them to visit a healthcare provider’s office or the hospital. Before the COVID-19 pandemic, virtual care was often used to provide medical care to patients from rural and distant communities; however, it is now being used routinely in urban settings.^1^ Videoconference assessments (e.g. Zoom) may offer further advantages over telephone-based assessments because healthcare providers gain additional information from some components of the physical (e.g. gait assessment) and mental status examinations that are not feasible by telephone.^2^

Although virtual care is a way for healthcare providers to continue caring for patients throughout the COVID-19 pandemic, certain factors (e.g. cognitive impairment, clinical frailty) and social disparities (e.g. social isolation, completion of less formal education) could prevent patients from accessing videoconference-based virtual care.^4^ Participating in videoconference assessments requires both access to and knowledge of web-based technology (e.g. Zoom). Our objective was to identify patient-specific factors associated with accessing videoconference assessments, as opposed to telephone-based assessments, in a diverse population of older adults. Our findings will inform future initiatives supporting older adults to better access videoconference-based virtual care.

## Methods

This manuscript is reported in accordance with the STROBE (Strengthening the Reporting of Observational Studies in Epidemiology) statement for the reporting of observational studies.^5^ We obtained ethics approval for this study from the Unity Health Toronto Research Ethics Board.

### Setting and Data Source

We retrospectively reviewed the medical records of all patients who received at least one virtual assessment in the geriatric medicine clinic at St. Michael’s Hospital-Unity Health Toronto, Toronto, Canada, between March 17, 2020, and July 13, 2020. March 17, 2020 was the first day that patients were seen by telephone or videoconference, as opposed to in-person, during the COVID-19 pandemic. The mode of virtual assessment was chosen by patients or their caregivers. The geriatric medicine clinic at St. Michael’s Hospital serves a diverse inner-city population of older adults. All study data were extracted from patients’ comprehensive geriatric assessments and follow-up clinic notes. Many patients in our study population were seen in follow-up, but were initially referred to the geriatric medicine clinic through primary care, inpatient, or emergency department settings.

### Study Design

We implemented a retrospective cross-sectional study design. We extracted the following data from each patient’s medical record: age, sex, modality of virtual care (i.e. telephone or videoconference), caregiver involvement in assessment (present or absent), language of assessment, highest level of education completed (e.g. elementary school, high school, post-secondary), birthplace (i.e. in Canada or another country), cognitive status (i.e. presence or absence of cognitive impairment), independence in completing basic activities of daily living (i.e. bathing, eating, ambulating, toileting, and hygiene), independence in completing instrumental activities of daily living (i.e. shopping, housework, accounting, food preparation, transportation, medication administration, and telephone and computer usage), symptomatic disease (e.g. dyspnea from chronic obstructive pulmonary disease), and frailty. We described frailty status with the Clinical Frailty Scale (CFS), which captures a patient’s ability to withstand an acute stressor. CFS scores range from 1 (very fit) to 9 (terminally ill).^6^ Where the CFS score was not explicitly stated by the treating geriatrician, patients were assigned a CFS score (by JAW [a geriatrician experienced in administering the CFS]) based on the described burden of symptomatic disease and impairments in basic and instrumental activities of daily living. Patients were classified as having cognitive impairment if the treating geriatrician described a diagnosis of mild cognitive impairment, dementia, or vascular cognitive impairment.^7, 8^

### Statistical Analysis

We reported the median patient age (and interquartile range [IQR]) and mean CFS score (and standard deviation [SD]), by assessment group (i.e. telephone or videoconference). We compared group age distributions with a Wilcoxon rank-sum test and group mean CFS scores with a pooled Student’s t-test. All other variables were analyzed as categorical variables, which were summarized as frequencies and percentages and compared across groups with chi square tests or Fisher’s exact tests when expected cell frequencies were five or less.

We used a multivariable logistic regression model to test the association between having a videoconference assessment (vs. a telephone assessment) and patient age, sex (male vs. female), education (post-secondary vs. high school equivalent or less), CFS score, history of cognitive impairment (cognitive impairment vs. no cognitive impairment), immigration history (born in vs. immigrated to Canada), language of assessment (all languages other than English vs. English), caregiver involvement (absent vs. present), and ability to use a computer (independent vs. dependent). Based on our clinical experience and a review of the e-health literacy literature, we believed these patient-specific factors could impact patients’ ability to access videoconference as opposed to telephone-based virtual care.^9, 10^ We included missing values for categorical variables as an additional category. There were no missing values for continuous variables. We presented associations as unadjusted odds ratios (ORs), adjusted ORs, and adjusted risk differences (RDs) with 95% confidence intervals (CIs). We presented the following by caregiver presence or absence, (1) marginal probabilities of virtual assessment at representative values of the CFS (i.e. 3, 4, 5, 6, and 7) while allowing other variables to take observed sample values; and (2) predicted probabilities for an 80-year-old woman with CFS score 5 who immigrated to Canada, speaks English, attained a post-secondary education, does not have cognitive impairment, and independently uses a computer.^11, 12 13^ We reported two-sided p-values and considered p-values <0.05 as statistically significant. We conducted analyses in STATA, version 15.1.

## Results

We reviewed the medical records of 332 patients who were assessed virtually in the geriatric medicine clinic at St. Michael’s Hospital. Two patients were excluded because they had delirium at the time of assessment and we could not determine whether they had underlying cognitive impairment. Therefore, we included 330 patients in our study population. Among patients receiving a telephone-based assessment (number[n]=227), the median age was 83 (Q1-Q3 76 to 88), 46.26% (n=105) were male, and caregivers were absent for 44.93% (n=102) of assessments (Table 1). Among patients receiving a virtual assessment (n=103), the median age was 84 (Q1-Q3 77 to 87), 42.72% (n=44) were male, and caregivers were absent for 44.93% (n=102) of assessments (Table 1). Only 32 of 98 (32.7%) patients who could independently use a computer participated in videoconference assessments.

**Table 1.** Baseline characteristics of older adults receiving a telephone or videoconference assessment

|  | Telephone<br>(n=227) | Videoconference<br>(n=103) | p-value |
| --- | --- | --- | --- |
| Age (years), median (Q1 to Q3) | 83 (76 to 88) | 84 (77 to 87) | 0.70 |
| Male sex, n (%) | 105 (46.3) | 44 (42.7) | 0.55 |
| Frailty (Clinical Frailty Scale), mean (SD) | 5.1 (1.0) | 4.9 (0.9) | 0.14 |
| Caregiver absent at assessment, n (%) | 102 (44.9) | 15 (14.6) | <0.01 |
| Assessment in English, n (%) | 198 (87.2) | 89 (86.4) | 0.84 |
| Cognitive impairment, n (%) | 186 (81.9) | 78 (75.7) | 0.19 |
| Highest level of education completed |  |  |  |
| Post-secondary, n (%) | 137 (60.4) | 58 (56.3) | 0.77 |
| High school or less, n (%) | 83 (36.6) | 42 (40.8) |  |
| Missing, n (%) | 7 (3.1) | 3 (2.9) |  |
| Immigration status |  |  |  |
| Born in Canada, n (%) | 93 (41.0) | 41 (39.8) | 0.36 |
| Immigrated to Canada, n (%) | 131 (57.7) | 58 (56.3) |  |
| Missing, n (%) | 3 (1.3) | 4 (3.9) |  |
| Independence in using a computer |  |  |  |
| Yes, n (%) | 66 (29.1) | 32 (31.1) | 0.89 |
| No, n (%) | 42 (18.5) | 20 (19.4) |  |
| Missing, n (%) | 119 (52.4) | 51 (49.5) |  |

Abbreviations: first quartile (Q1, third quartile (Q3), number (n), standard deviation (SD)

Patients with frailty (adjusted OR 0.62, 0.45 to 0.85; adjusted RD -0.08, -0.09 to -0.06) or who lacked a caregiver to facilitate a virtual assessment (adjusted OR 0.12, 0.06 to 0.24; adjusted RD -0.35, -0.43 to -0.26) had significantly lower odds of receiving a videoconference compared to telephone assessment (Table 2). There were no significant differences in age, sex, level of education, immigration status, history of cognitive impairment, language of assessment, or ability to use a computer between videoconference and telephone assessment groups (Table 2). Data pertaining to independence in using a computer were missing for 52.42% (n=119) of persons receiving a telephone assessment and 49.51% (n=51) of persons receiving a videoconference assessment (Table 1).

**Table 2.**
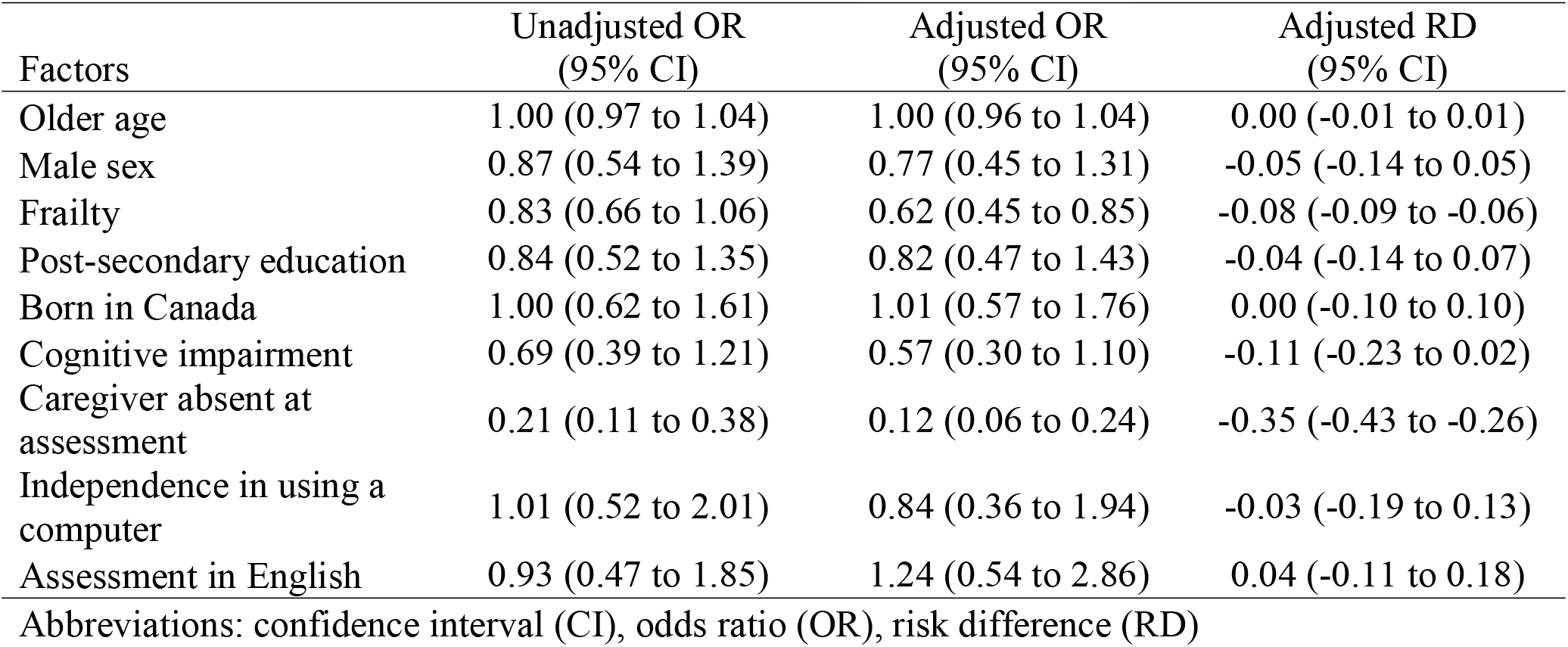
Factors associated with videoconference versus telephone assessment

| Factors | Unadjusted OR<br>(95% CI) | Adjusted OR<br>(95% CI) | Adjusted RD<br>(95% CI) |
| --- | --- | --- | --- |
| Older age | 1.00 (0.97 to 1.04) | 1.00 (0.96 to 1.04) | 0.00 (-0.01 to 0.01) |
| Male sex | 0.87 (0.54 to 1.39) | 0.77 (0.45 to 1.31) | -0.05 (-0.14 to 0.05) |
| Frailty | 0.83 (0.66 to 1.06) | 0.62 (0.45 to 0.85) | -0.08 (-0.09 to -0.06) |
| Post-secondary education | 0.84 (0.52 to 1.35) | 0.82 (0.47 to 1.43) | -0.04 (-0.14 to 0.07) |
| Born in Canada | 1.00 (0.62 to 1.61) | 1.01 (0.57 to 1.76) | 0.00 (-0.10 to 0.10) |
| Cognitive impairment | 0.69 (0.39 to 1.21) | 0.57 (0.30 to 1.10) | -0.11 (-0.23 to 0.02) |
| Caregiver absent at assessment | 0.21 (0.11 to 0.38) | 0.12 (0.06 to 0.24) | -0.35 (-0.43 to -0.26) |
| Independence in using a computer | 1.01 (0.52 to 2.01) | 0.84 (0.36 to 1.94) | -0.03 (-0.19 to 0.13) |
| Assessment in English | 0.93 (0.47 to 1.85) | 1.24 (0.54 to 2.86) | 0.04 (-0.11 to 0.18) |

Abbreviations: confidence interval (CI), odds ratio (OR), risk difference (RD)

If a patient did not have a caregiver present to facilitate a virtual assessment, the predicted probability of receiving a videoconference compared to telephone assessment was less than 50%, regardless of frailty status, ability to use a computer, or history of cognitive impairment (Figure and Supplementary Figures S1 and S2). The predicted probability of receiving a virtual compared to telephone assessment for an 80-year-old woman with a CFS score of 5 who immigrated to Canada, speaks English, attained a post-secondary education, does not have cognitive impairment, and independently uses a computer was 60% (39% to 80%) if a caregiver could facilitate a virtual assessment; however, her predicted probability of receiving a virtual assessment decreased to 15% (3% to 26%) if a caregiver could not facilitate a virtual assessment.

**Figure.**
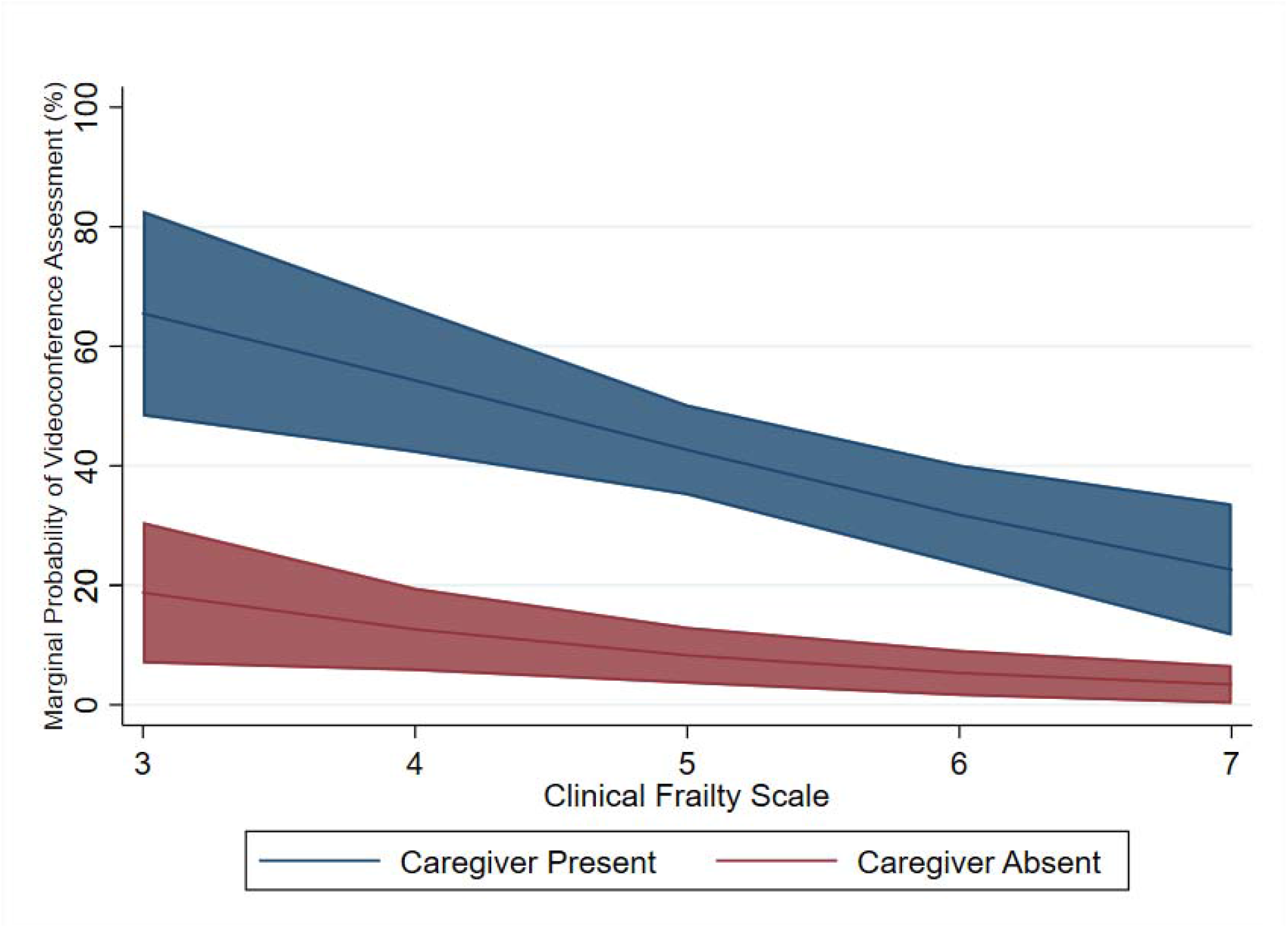
Marginal probability (and 95% confidence region) of receiving a videoconference compared to telephone assessment, by frailty status and caregiver presence or absence.

## Discussion

In this retrospective cross-sectional study of older adults assessed virtually (i.e. by videoconference or telephone) in a geriatric medicine clinic during the COVID-19 pandemic, we found that frailty and the absence of a caregiver to facilitate virtual assessments were associated with significantly lower odds of receiving a videoconference compared to telephone-based assessment. Our results suggest that clinicians, researchers, and policymakers must urgently implement and evaluate strategies that optimise equitable access to videoconference-based virtual care for older adults.

Our results build upon a growing literature suggesting that (1) older adults may be less likely to access videoconference compared to a telephone assessments and (2) factors unique to older adults might be important in understanding barriers to accessing videoconference-based virtual care.^14^ Within the Oxford Royal College of General Practitioners (RCGP) Research and Surveillance Centre, 55.1% of geriatric medicine appointments occurred over the telephone by week 14 of the COVID-19 pandemic; whereas, only 1.5% of these appointments occurred by videoconference.^15^ In Northern Finland, a population-based survey of older adults conducted before the COVID-19 pandemic found that frail older adults were less likely to use the internet or other communication technologies, including Skype, suggesting that frailty might not be contributing to this lower use of videoconference assessments.^4^ Specifically, there was an inverse association between frailty and having an internet connection at home (46.4% of frail older adults compared to 78.6% of non-frail older adults, *P* < 0.001).^4^ And, frail older adults were less likely to have used a computer in the past 12 months (30.4% of frail older adults compared to 70.0% non-frail older adults, *P* < 0.001).^4^ In the United States, findings from the 2018 National Health and Aging Trends Study suggested that 38% of older adults were not ready for videoconference assessments, predominantly because of their inexperience with technology.^14^ Using videoconferencing requires both access to and knowledge of web-based technology (e.g.Zoom). Therefore, even if older adults have access to a computer, which they may use regularly for tasks such as checking email, they may not be familiar with how to use the web-based technology needed to access videoconference-based medical care.

Although the COVID-19 pandemic accelerated adoption of virtual care, it was already carving out a place in healthcare, which further drives the need to ensure that older adults have equitable access to both modes of virtual care.^3, 16^ Future research should focus on demonstrating how caregivers and healthcare providers can support older adults in accessing videoconference-based virtual care; comparing the quality of care that patients receive when assessments are completed by telephone, videoconference, or in-person; and incorporating independence in computer use as an instrumental activity of daily living in assessments.

Our study had limitations. Data describing independence in using a computer were missing for 51.5% of patients. However, the proportion of missing data across virtual assessment groups (i.e. telephone or videoconference) was balanced and we accounted for these missing data in our analysis by creating an additional category for missing data (i.e. independent, dependent, or unknown). Furthermore, our study did not focus on patients who could not access virtual assessments; therefore, our study does not describe other potentially important factors such as hearing impairment, which could make any form of virtual assessment more difficult.

As the COVID-19 pandemic evolves, we need to better support older adults in accessing videoconference-based virtual care. We identified actionable solutions that can be implemented in routine practice: identify frailty and inquire about a caregiver who can facilitate virtual care. Our results do not necessarily mean that treating frailty will improve access to videoconference-based virtual care. However, in addition to our findings, there are many reasons why we should identify and treat frailty to slow its progression. Frailty is associated with adverse outcomes (e.g. increased risk of death from COVID-19, hospitalization, nursing home admission, and adverse drug events) and it is treatable with multicomponent interventions (e.g. enhanced nutrition, cognitive stimulation, and physical activity).^17-20^ Furthermore, our findings suggest that older adults may benefit from a caregiver’s support to facilitate access to videoconference-based assessments, paving the way for initiatives that increase the involvement of patients’ loved ones or volunteers. Future research should aim to implement and evaluate interventions that support older adults in being part of the digital healthcare revolution.

## Supporting information

Reporting Checklist

## Data Availability

Interested researchers should contact the corresponding author for further information.

## Author Contributions

ZG, AJ, RP, SES, and JAW designed the study. LL and JW completed all data abstraction. JAW completed all data analysis. LL and JAW drafted the manuscript. LL, ZG, AJ, RP, SES, and JAW edited the manuscript. All authors read and approved the final manuscript prior to its submission. LL and JAW had full access to all of the data (including statistical reports and tables) in the study and can take responsibility for the integrity of the data. JAW takes responsibility for the accuracy of the data analysis and affirms that the manuscript is an honest, accurate, and transparent account of the study being reported.

## References

[1] Hardcastle L, Ogbogu U. Virtual care: Enhancing access or harming care? Healthc Manage Forum. 2020: 840470420938818.

[2] Wosik J, Fudim M, Cameron B, et al. Telehealth transformation: COVID-19 and the rise of virtual care. J Am Med Inform Assoc. 2020;27: 957–962.

[3] Force VCT. Virtual Care: Recommendations for Scaling Up Virtual Medical Services. In: Association CM, ed., 2020.

[4] Keranen NS, Kangas M, Immonen M, et al. Use of Information and Communication Technologies Among Older People With and Without Frailty: A Population-Based Survey. J Med Internet Res. 2017;19: e29.

[5] von Elm E, Altman DG, Egger M, et al. The Strengthening the Reporting of Observational Studies in Epidemiology (STROBE) Statement: guidelines for reporting observational studies. Int J Surg. 2014;12: 1495–1499.

[6] Rockwood K, Song X, MacKnight C, et al. A global clinical measure of fitness and frailty in elderly people. CMAJ. 2005;173: 489–495.

[7] Diagnostic and Statistical Manual of Mental Disorders. 5th ed. Arlington, VA: American Psychiatric Association 2013.

[8] O’Brien JT, Erkinjuntti T, Reisberg B, et al. Vascular cognitive impairment. Lancet Neurology. 2003;2: 89–98.

[9] Roque NA, Boot WR. A New Tool for Assessing Mobile Device Proficiency in Older Adults: The Mobile Device Proficiency Questionnaire. J Appl Gerontol. 2018;37: 131–156.

[10] Sudbury-Riley L, FitzPatrick M, Schulz PJ. Exploring the Measurement Properties of the eHealth Literacy Scale (eHEALS) Among Baby Boomers: A Multinational Test of Measurement Invariance. J Med Internet Res. 2017;19: e53.

[11] Williams R. Using the margins command to estimate and interpret adjusted predictions and marginal effects. The Stata Journal. 2012;12: 308–331.

[12] Norton EC, Dowd BE, Maciejewski ML. Marginal Effects-Quantifying the Effect of Changes in Risk Factors in Logistic Regression Models. JAMA. 2019;321: 1304–1305.

[13] Hammer MJ, Kalkan KO. Behind the Curve: Clarifying the Best Approach to Calculating Predicted Probabilities and Marginal Effects from Limited Dependent Variable Models. American Journal of Political Science. 2012;57: 263–277.

[14] Lam K, Lu AD, Shi Y, Covinsky KE. Assessing Telemedicine Unreadiness Among Older Adults in the United States During the COVID-19 Pandemic. JAMA Intern Med. 2020.

[15] Joy M, McGagh D, Jones N, et al. Reorganisation of primary care for older adults during COVID-19: a cross-sectional database study in the UK. Br J Gen Pract. 2020.

[16] Shaw J, Jamieson T, Agarwal P, Griffin B, Wong I, Bhatia RS. Virtual care policy recommendations for patient-centred primary care: findings of a consensus policy dialogue using a nominal group technique. J Telemed Telecare. 2018;24: 608–615.

[17] Orgeta V, Leung P, Yates L, et al. Individual cognitive stimulation therapy for dementia: a clinical effectiveness and cost-effectiveness pragmatic, multicentre, randomised controlled trial. Health Technol Assess. 2015;19: 1–108.

[18] Hewitt J, Carter B, Vilches-Moraga A, et al. The effect of frailty on survival in patients with COVID-19 (COPE): a multicentre, European, observational cohort study. The Lancet Public Health. 2020.

[19] Fried LP, Tangen CM, Walston J, et al. Frailty in Older Adults: Evidence for a Phenotype. Journal of Gerontology: MEDICAL SCIENCES. 2001;56A: M146–M156.

[20] Maxwell CJ, Campitelli MA, Hogan DB, et al. Relevance of frailty to mortality associated with the use of antipsychotics among community-residing older adults with impaired cognition. Pharmacoepidemiol Drug Saf. 2018;27: 289–298.

