## Supplemental File for "Factors associated with access to virtual care in older adults: A cross-sectional study"

Supplementary File

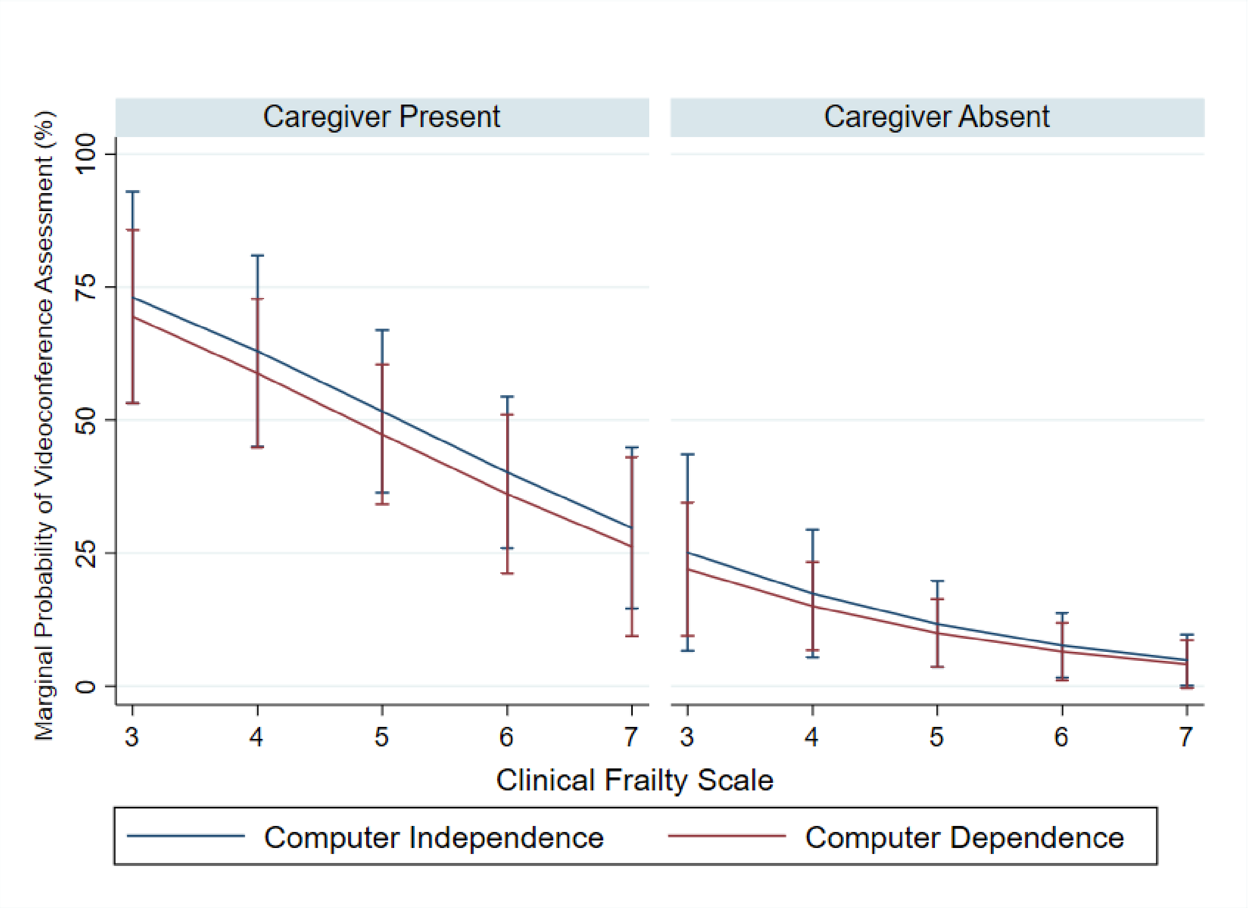


Supplementary Figure S1. Marginal probability (and 95% confidence interval) of videoconference assessment for study participants who were independent in using a computer compared to those who were not independent in using a computer, by caregiver presence or absence and Clinical Frailty Scale score

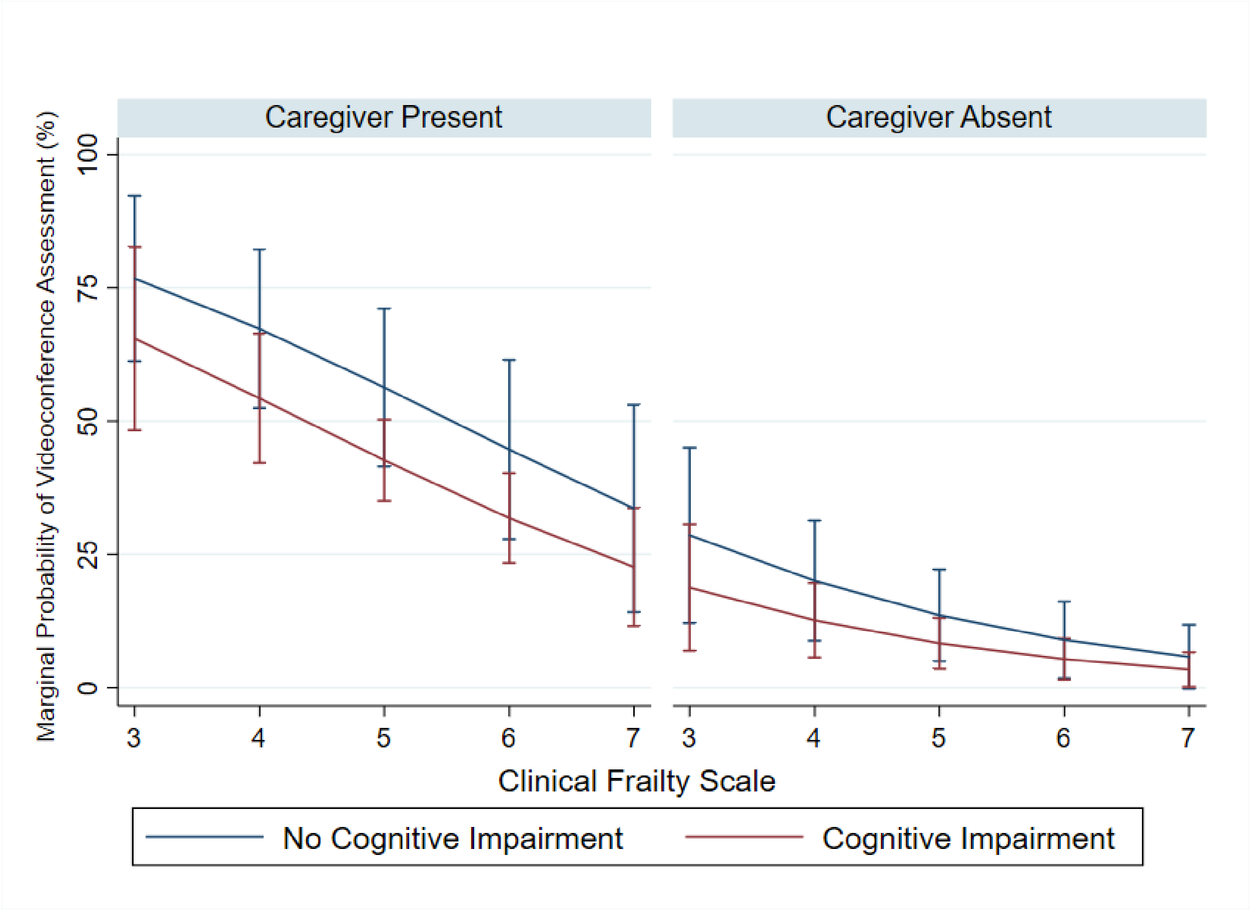


Supplementary Figure S2. Marginal probability (and 95% confidence interval) of videoconference assessment for study participants without cognitive impairment compared to those who had cognitive impairment, by caregiver presence or absence and Clinical Frailty Scale score
